# Assessment of adaptive functioning in Angelman syndrome using the Vineland Adaptive Behavior Scales, Third Edition

**DOI:** 10.64898/2026.06.11.26355399

**Authors:** Sarah Nelson Potter, Julia Zhang, Batsheva Friedman, Julia Gable, Nadia Ali, Rene L. Barbieri-Welge, Ayala Ben-Tall, Kelly E. Caravella, Margaret DeRamus, Dea Garic, Margot MacKay, Kara Murias, Sarika U. Peters, Kimberly Smyth, Jane Summers, Angel Wang, Mark D. Shen, Jörg F. Hipp, Julian Tillmann, Jorrit Tjeertes, Brenda Vincenzi, Lynne M. Bird, Wen-Hann Tan, Anne C. Wheeler, Anjali Sadhwani

## Abstract

**Purpose:** This study examined longitudinal trajectories of adaptive functioning in 331 individuals with Angelman syndrome (AS) using the Vineland Adaptive Behavior Scales, Third Edition (Vineland-3) and examined differences by molecular subtype.

**Methods:** A total of 331 individuals (156 females, 47%) with genetically confirmed AS (ages 6 months to 52 years) were assessed between 2018 and 2025, including 207 with a deletion subtype, 63 with uniparental disomy or imprinting defect, and 61 with a *UBE3A* point mutation. Growth scale values were analyzed using linear mixed-effects models with log_2_-transformed age.

**Results:** Individuals with deletion subtypes demonstrated significantly lower adaptive functioning across domains compared to those with non-deletion subtypes. Adaptive skills across all Vineland-3 subdomains increased nonlinearly with age, showing faster growth early in life that slowed over time, with largely parallel trajectories across subtypes.

**Conclusion:** Individuals with AS demonstrate slow but steady growth in adaptive functioning that continues into adulthood, with progress varying by molecular subtype. These findings provide updated natural history benchmarks and demonstrate the utility of the Vineland-3 for clinical trials.

---

Angelman syndrome (AS) is a rare neurogenetic condition resulting from the loss of expression of the maternally inherited copy of the ubiquitin-protein ligase E3A (*UBE3A*) gene on chromosome 15q11-q13 (Bird, 2014). This genetic alteration leads to a deficiency of UBE3A protein in neurons, resulting in a characteristic phenotype that includes severe to profound intellectual disability, minimal to absent speech, motor impairments, epilepsy, and a distinctive behavioral profile marked by frequent laughter, excitability, and a generally happy demeanor (Bird, 2014; Pearson et al., 2019). AS has four known genetic subtypes: deletion (affecting ∼65-70% of individuals), *UBE3A* point mutation (∼10-15%), imprinting defect (ImpD; ∼5-10%), and paternal uniparental disomy (UPD; ∼5-10%) (Dagli et al., 1993). These latter three subtypes are collectively referred to as *non-deletion* AS.

Adaptive functioning involves the conceptual, social, and practical skills that individuals use in their daily lives. Individuals with AS, regardless of molecular subtype, experience profound and persistent challenges in adaptive functioning, particularly in communication and daily living skills (Brun Gasca et al., 2010; Di Nuovo & Buono, 2011; Gentile et al., 2010; Gwaltney et al., 2024; Keute et al., 2021; Peters et al., 2004), resulting in impairments that necessitate lifelong care (Wheeler et al., 2017). A recent study examined adaptive skills and trajectories over time in individuals with AS between one and 13 years of age using the second edition of the Vineland Adaptive Behavior Scales (Vineland-II) (Gwaltney et al., 2024; Sparrow et al., 2005). In that study, 257 participants were assessed at one of six study sites between 2006 and 2014 as part of the AS Natural History Study (NCT00296764). Consistent with previous studies, Gwaltney and colleagues found that individuals with deletion subtypes demonstrated a lower level of adaptive skills relative to those with non-deletion subtypes (Brun Gasca et al., 2010; Di Nuovo & Buono, 2011; Gentile et al., 2010; Keute et al., 2021; Peters et al., 2004). Statistically significant growth was observed in all adaptive domains throughout childhood and into early adolescence. At six years of age, individuals with deletion subtype had age equivalent (AE) scores ranging from 12 to 18 months, whereas those with non-deletion subtypes had AE scores ranging from 18 to 38 months in all subdomains except Expressive Communication. Regardless of molecular subtype, individuals with AS demonstrated a relative weakness on the Expressive Communication subdomain, with AE scores ranging from approximately 10 to 14 months at six years of age.

An updated version of the Vineland (i.e. Vineland-3) was released in 2016, with several significant changes that are important for assessing individuals with intellectual and developmental disabilities (IDD) (Sparrow et al., 2016). For example, on the newer measure, the lower bounds of the AE score ranges were modified for the Written, Domestic, Community, and Coping Skills subdomains (minimum AD scores of <3:0 for Written, Domestic, and Community subdomains; <2:0 for Coping Skills subdomain) to better reflect developmental expectations within these areas of adaptive functioning. The previous AE scores associated with a raw score of 0 were 1:10 for Written, 0:7 for Domestic, and <0:1 for Community and Coping Skills. An increased number of items assessing early skills on several subdomains (i.e., Receptive Communication, Written Communication, Personal Skills, Community Skills) as well as updated scoring criteria (Farmer et al., 2020) may help reduce floor effects that have been observed in previous studies, including Gwaltney et al. (2024). One particularly notable improvement on the Vineland-3 that is particularly relevant for individuals with AS is the inclusion of augmentative and alternative communication (AAC) modalities in the Communication domain, which were not accounted for in the Vineland-II. Given the significant deficits in spoken communication observed in most individuals with AS, and the number of individuals who rely on AAC modalities (e.g., gestures, sign language, pictures, switches or buttons, speech-generating devices) (Pearson et al., 2019), the inclusion of AAC in the Vineland-3 is a significant improvement for more accurately measuring communication skills in AS.

In addition to the substantial changes between the Vineland-II and Vineland-3, the Vineland-3 is included as a core secondary clinical outcome assessment measure in several recent and upcoming AS clinical trials (e.g., NCT04428281, NCT05011851, NCT05127226, NCT06415344, NCT06617429, NCT06914609, NCT07181837), underscoring the importance of updating the natural history of adaptive functioning in AS using the most up-to-date version of the Vineland. Specifically, we sought to: (1) characterize longitudinal trajectories of skills across the Vineland-3 domains of Communication, Daily Living Skills, Socialization, and Motor Skills, and (2) describe differences in adaptive functioning across these domains by molecular subtype.

## Methods

### Participants

The sample consisted of 331 individuals with a molecular diagnosis of AS confirmed via review of clinical diagnostic reports. Between 2018 and 2025, data for these participants were collected longitudinally at the UNC Comprehensive Angelman Syndrome Clinic, or as part of the AS Natural History Study (ClinicalTrials.gov identifier: NCT04507997) or the FREESIAS study (Tjeertes et al., 2023). These studies were conducted at the following sites: Alberta Children’s Hospital, Boston Children’s Hospital, British Columbia Children’s Hospital, Carolina Institute for Developmental Disabilities, Children’s Hospital Colorado, Children’s Hospital of Eastern Ontario, Children’s Hospital Los Angeles, Emory University Hospital, Rady Children’s Hospital-San Diego, Rush University Medical Center, Texas Children’s Hospital, and Vanderbilt University Medical Center. Vineland interviews were administered with a primary caregiver and conducted or supervised by a licensed psychologist with expertise in AS. Participants were excluded from analyses if they were known to have mosaicism. Participants between the ages of 6 months and 52 years were included in analyses.

### Measures

The Vineland-3 is a norm-referenced standardized assessment that captures adaptive skills from birth through 90 years of age. The Comprehensive Interview Form of the Vineland-3 was used to assess adaptive functioning in the domains of Communication, Daily Living Skills, Socialization, and Motor Skills. The Communication domain assesses receptive, expressive, and written skills. The Written subdomain, however, was not included in the current study due to the limited number of individuals with AS who demonstrate skills represented in this subdomain. The Daily Living Skills domain measures skills in the areas of personal, domestic, and community functioning. The Socialization domain examines interpersonal relationships, play and leisure, and coping skills. Finally, the Motor Skills domain evaluates both gross and fine motor abilities.

Multiple scoring options are available for the Vineland-3. The publisher (i.e., Pearson) provides standard scores for each domain and the overall Adaptive Behavior Composite (mean = 100, SD = 15) as well as subdomain standard scores (i.e., v-scale scores; mean = 15, SD = 3). However, age-normed standardized scores are often not sufficient for capturing the full range of functioning in individuals with IDD, including those with AS, due to floor effects (Thurm et al., 2020). For this reason, we examined alternative scoring options, namely AE scores and growth scale values (GSVs). Both AE scores and GSVs are derived from raw scores, with GSVs offering the advantage of being on an equal-interval scale. Because a one-unit change on the GSV scale carries the same meaning across the entire range, GSVs are more sensitive than other scoring options for detecting small changes in adaptive functioning over time. This feature makes them particularly useful for modeling within-person change over time (Eisengart et al., 2022; Farmer et al., 2025; Farmer et al., 2023; Kwok et al., 2022; Psimas & Williams, 2024). Importantly, GSVs are not comparable across subdomains; a one-unit change on one subdomain does not represent the same level of change on another subdomain. Accordingly, all analyses and interpretations of GSVs in the current study were conducted within subdomains rather than across subdomains. Notably, AE scores are less useful than GSVs for tracking change over time because a range of raw scores can correspond to the same AE value.

### Data Analysis Plan

Baseline participant characteristics and AE scores were compared across three molecular subtype groups (i.e., deletion, *UBE3A* mutation, UPD/ImpD) as well as between each non-deletion group and the deletion group. Individuals with UPD and imprinting defect functionally have two paternal copies of the chromosome 15q11q13 region, instead of one from each parent. Moreover, previous studies have shown that these individuals have similar neurodevelopmental profiles and medical complications (Keute et al., 2021). Given the relatively small number of participants in each of these subgroups, they were combined for the purposes of our analyses. Categorical variables were summarized by frequencies with percentages and compared using Pearson chi-squared tests or Fisher’s exact tests when cell counts were less than five. Continuous variables were summarized using the mean and standard deviation and compared using the Kruskal-Wallis test for three group comparisons and the Wilcoxon rank-sum test for post hoc two-group comparisons (i.e., deletion vs. *UBE3A* mutation, deletion vs. UPD/ImpD, *UBE3A* mutation vs. UPD/ImpD).

To compare associations between subdomain GSVs and age across molecular subtypes, a linear mixed model (LMM) with restricted maximum likelihood was fitted for each Vineland-3 subdomain. Each model included a random effect for participant to account for within-subject correlation due to repeated longitudinal measurements. Fixed effects in each LMM included molecular subtype, age, and the interaction of molecular subtype with age, allowing the association between age and GSVs to differ by molecular subtype. Given the nonlinear relationship between age and GSVs, a logarithmic transformation of age was selected after comparison with alternative models (e.g., linear and quadratic age terms and logarithmic outcome transformation) using Akaike Information Criterion and Bayesian Information Criterion and the marginal R^2^. See supplemental material for a comparison of model metrics for the Receptive Communication subdomain as an example. For easier interpretation of the main effect of age, age was transformed using a base-2 logarithm and centered at log_2_(11), where 11 years represents the mean age of participants in the reference group (i.e., deletion). In these models, the main effect of age represented the average change in GSVs associated with a doubling of age for individuals in the reference group. Equivalent analyses using raw scores are available in supplemental material.

All *p-*values were two-sided, and statistical significance was defined as *p* < .05. The widths of the 95% confidence intervals were not adjusted for multiple comparisons. Analyses were performed using SAS® software (version 9.4, SAS Institute, Cary, North Carolina) and R (version 4.5.1, R Core Team 2023).

## Results

### Sample Characteristics

The final sample was relatively evenly split between females (*n* = 156, 47%) and males (*n* = 175, 53%). Approximately 63% of participants had a deletion, 18% had a *UBE3A* mutation, and 19% had UPD or ImpD. Participants had a mean age of 11.3 years (*SD* = 10.0) at baseline, with no significant differences across molecular subtypes. The number of study visits ranged from one to six, with a mean of two visits. Additional participant characteristics are summarized in **Table 1**.

**Table 1.** Participant Characteristics by Molecular Diagnosis.

| **Variable** | **Deletion**<br>**(n = 207)** | **UPD/ImpD**<br>**(n = 63)** | **UBE3A**<br>**mutation**<br>**(n = 61)** | **Overall**<br>**(N = 331)** | **p-value** |
| --- | --- | --- | --- | --- | --- |
| **Sex, n (%)** |  |  |  |  | .193 |
| Female | 99 (48%) | 33 (54%) | 24 (38%) | 156 (47%) |  |
| Male | 108 (52%) | 28 (46%) | 39 (62%) | 175 (53%) |  |
| **Race<sup>1</sup>, n (%)** |  |  |  |  | .138 |
| White | 117 (57%) | 49 (80%) | 46 (73%) | 212 (64%) |  |
| Black | 6 (3%) | 3 (5%) | 2 (3%) | 11 (3%) |  |
| Asian | 16 (8%) | 1 (2%) |  | 17 (5%) |  |
| Other | 3 (1%) |  | 1 (2%) | 4 (1%) |  |
| Multiracial | 15 (7%) | 5 (8%) | 5 (8%) | 25 (8%) |  |
| Unknown | 50 (24%) | 3 (5%) | 9 (14%) | 62 (19%) |  |
| **Ethnicity**<br>**(Hispanic/Latino), n (%)** |  |  |  |  | .005 |
| Yes | 34 (16%) | 5 (8%) | 7 (11%) | 46 (14%) |  |
| No | 122 (59%) | 51 (84%) | 46 (73%) | 219 (66%) |  |
| Unknown | 51 (25%) | 5 (8%) | 10 (16%) | 66 (20%) |  |
| **Age at Baseline (Years)** |  |  |  |  | .911 |
| Mean (SD) | 11.4 (10.5) | 10.9 (8.5) | 11.3 (9.6) | 11.3 (10.0) |  |
| Range | 0.5 - 52.1 | 1.3 - 34.0 | 0.8 - 37.9 | 0.5 - 52.1 |  |
| **Age at Last Visit (Years)** |  |  |  |  | .773 |
| Mean (SD) | 12.8 (10.9) | 12.3 (8.3) | 12.8 (9.8) | 12.7 (10.2) |  |
| Range | 1.3 - 52.1 | 1.7 - 34.0 | 1.2 - 42.8 | 1.2 - 52.1 |  |
| **Number of Visits** |  |  |  |  | .349 |
| Mean (SD) | 1.9 (1.1) | 2.1 (1.1) | 2.0 (1.3) | 2.0 (1.1) |  |
| Range | 1 - 5 | 1 - 6 | 1 - 6 | 1 - 6 |  |

*Note.* Continuous variables were compared using Kruskal-Wallis tests. Categorical variables were compared using Pearson chi-square tests. <sup>1</sup>Fisher's exact test was used due to small cell counts (<5) in multiple cells. UPD: uniparental disomy. ImpD: imprinting defect.

### Age Equivalent Scores at Baseline Visit

**Table 2** displays mean baseline AE scores (in months) for each Vineland-3 subdomain across molecular subtypes. Kruskal-Wallis tests indicated significant differences across all subdomains (*p*’s < .001), with the deletion group showing consistently lower AE scores on average compared with the non-deletion groups. Post hoc comparisons between the non-deletion groups using Wilcoxon rank sum tests only showed significant differences on the Interpersonal Relationships (*p* = .018) and Play and Leisure (*p* = .004) subdomains, with those in the *UBE3A* mutation group demonstrating greater AE scores compared to those in the UPD/ImpD group.

**Table 2.**
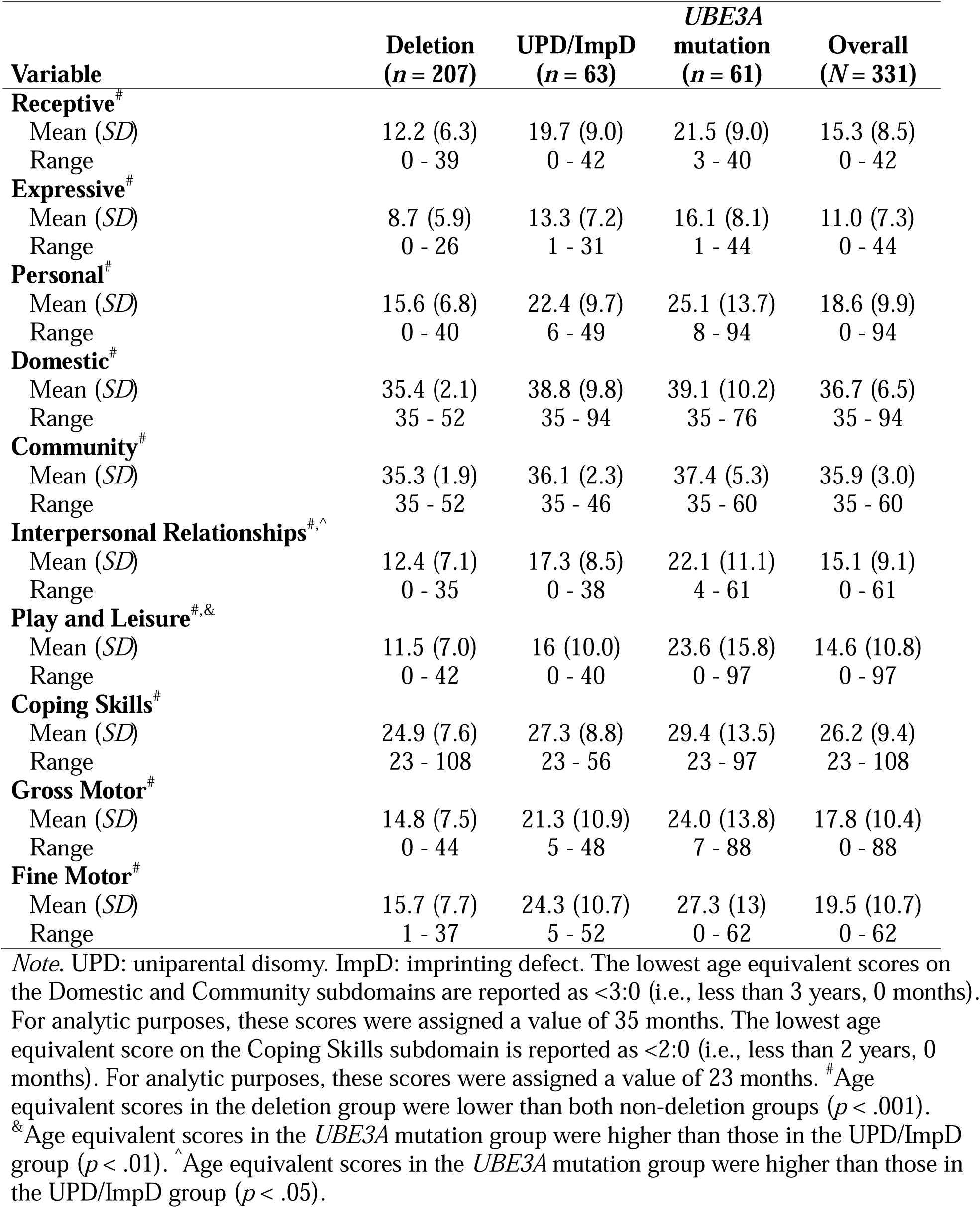
Baseline Vineland-3 Age Equivalent Scores (in Months) by Molecular Diagnosis Subtype.

| **Variable** | **Deletion**<br>**(n = 207)** | **UPD/ImpD**<br>**(n = 63)** | **UBE3A**<br>**mutation**<br>**(n = 61)** | **Overall**<br>**(N = 331)** |
| --- | --- | --- | --- | --- |
| **Receptive<sup>#</sup>** |  |  |  |  |
| Mean (SD) | 12.2 (6.3) | 19.7 (9.0) | 21.5 (9.0) | 15.3 (8.5) |
| Range | 0 - 39 | 0 - 42 | 3 - 40 | 0 - 42 |
| **Expressive<sup>#</sup>** |  |  |  |  |
| Mean (SD) | 8.7 (5.9) | 13.3 (7.2) | 16.1 (8.1) | 11.0 (7.3) |
| Range | 0 - 26 | 1 - 31 | 1 - 44 | 0 - 44 |
| **Personal<sup>#</sup>** |  |  |  |  |
| Mean (SD) | 15.6 (6.8) | 22.4 (9.7) | 25.1 (13.7) | 18.6 (9.9) |
| Range | 0 - 40 | 6 - 49 | 8 - 94 | 0 - 94 |
| **Domestic<sup>#</sup>** |  |  |  |  |
| Mean (SD) | 35.4 (2.1) | 38.8 (9.8) | 39.1 (10.2) | 36.7 (6.5) |
| Range | 35 - 52 | 35 - 94 | 35 - 76 | 35 - 94 |
| **Community<sup>#</sup>** |  |  |  |  |
| Mean (SD) | 35.3 (1.9) | 36.1 (2.3) | 37.4 (5.3) | 35.9 (3.0) |
| Range | 35 - 52 | 35 - 46 | 35 - 60 | 35 - 60 |
| **Interpersonal Relationships<sup>#,^</sup>** |  |  |  |  |
| Mean (SD) | 12.4 (7.1) | 17.3 (8.5) | 22.1 (11.1) | 15.1 (9.1) |
| Range | 0 - 35 | 0 - 38 | 4 - 61 | 0 - 61 |
| **Play and Leisure<sup>#,&</sup>** |  |  |  |  |
| Mean (SD) | 11.5 (7.0) | 16 (10.0) | 23.6 (15.8) | 14.6 (10.8) |
| Range | 0 - 42 | 0 - 40 | 0 - 97 | 0 - 97 |
| **Coping Skills<sup>#</sup>** |  |  |  |  |
| Mean (SD) | 24.9 (7.6) | 27.3 (8.8) | 29.4 (13.5) | 26.2 (9.4) |
| Range | 23 - 108 | 23 - 56 | 23 - 97 | 23 - 108 |
| **Gross Motor<sup>#</sup>** |  |  |  |  |
| Mean (SD) | 14.8 (7.5) | 21.3 (10.9) | 24.0 (13.8) | 17.8 (10.4) |
| Range | 0 - 44 | 5 - 48 | 7 - 88 | 0 - 88 |
| **Fine Motor<sup>#</sup>** |  |  |  |  |
| Mean (SD) | 15.7 (7.7) | 24.3 (10.7) | 27.3 (13) | 19.5 (10.7) |
| Range | 1 - 37 | 5 - 52 | 0 - 62 | 0 - 62 |

*Note.* UPD: uniparental disomy. ImpD: imprinting defect. The lowest age equivalent scores on the Domestic and Community subdomains are reported as <3:0 (i.e., less than 3 years, 0 months). For analytic purposes, these scores were assigned a value of 35 months. The lowest age equivalent score on the Coping Skills subdomain is reported as <2:0 (i.e., less than 2 years, 0 months). For analytic purposes, these scores were assigned a value of 23 months. <sup>#</sup>Age equivalent scores in the deletion group were lower than both non-deletion groups ( $p < .001$ ). <sup>&</sup>Age equivalent scores in the UBE3A mutation group were higher than those in the UPD/ImpD group ( $p < .01$ ). <sup>^</sup>Age equivalent scores in the UBE3A mutation group were higher than those in the UPD/ImpD group ( $p < .05$ ).

### Developmental Trajectories by Domain Across Molecular Subtypes

**Tables 3 to 6** present the model estimates for Vineland-3 GSVs across the Communication, Daily Living Skills, Socialization, and Motor Skills subdomains, respectively. **Figures 1 to 4** display the results of these models via effects plots, illustrating estimated GSV trajectories across age for each molecular subtype group. In these models, the intercept represents the predicted GSV for the deletion group at 11 years of age. Fixed-effects coefficients for the *UBE3A* mutation and UPD/ImpD groups indicate the predicted differences for these subtypes at age 11 relative to the deletion group (i.e., the reference group). The coefficient for age reflects the increase in GSVs with the doubling of age for the deletion group. Because age was log_2_-transformed, a positive coefficient indicates more rapid increases in scores at younger ages, with the rate of growth progressively slowing over time. Age-by-subtype interaction coefficients indicate how the effect of age differs for each non-deletion group relative to the deletion group.

#### Communication

For Receptive Communication, both non-deletion subtypes showed significantly higher GSVs than the deletion group at age 11 (deletion group GSV at age 11: 80.66). The *UBE3A* mutation group scored, on average, 21.75 points higher for a GSV score of 102.41 (*SE* = 2.23, *p* < .001), and the UPD/ImpD group scored 16.70 points higher with a GSV score of 97.36 (*SE* = 2.21, *p* < .001). The *UBE3A* mutation group and UPD/ImpD group did not differ significantly from each other at age 11. Age was positively associated with Receptive Communication GSVs, indicating that GSVs increased with age. From age 11 to 22, representing a doubling of age on the log_2_-transformed scale centered at 11 years, GSVs increased by 7.45 points in the deletion group, 8.15 points in the *UBE3A* mutation group, and 8.82 points in the UPD/ImpD group. The age-by-subtype interactions were not significant for either the *UBE3A* mutation or UPD/ImpD groups, indicating similar age-related trajectories across subtypes (**Table 3**, **Figure 1**).

**Figure 1.**
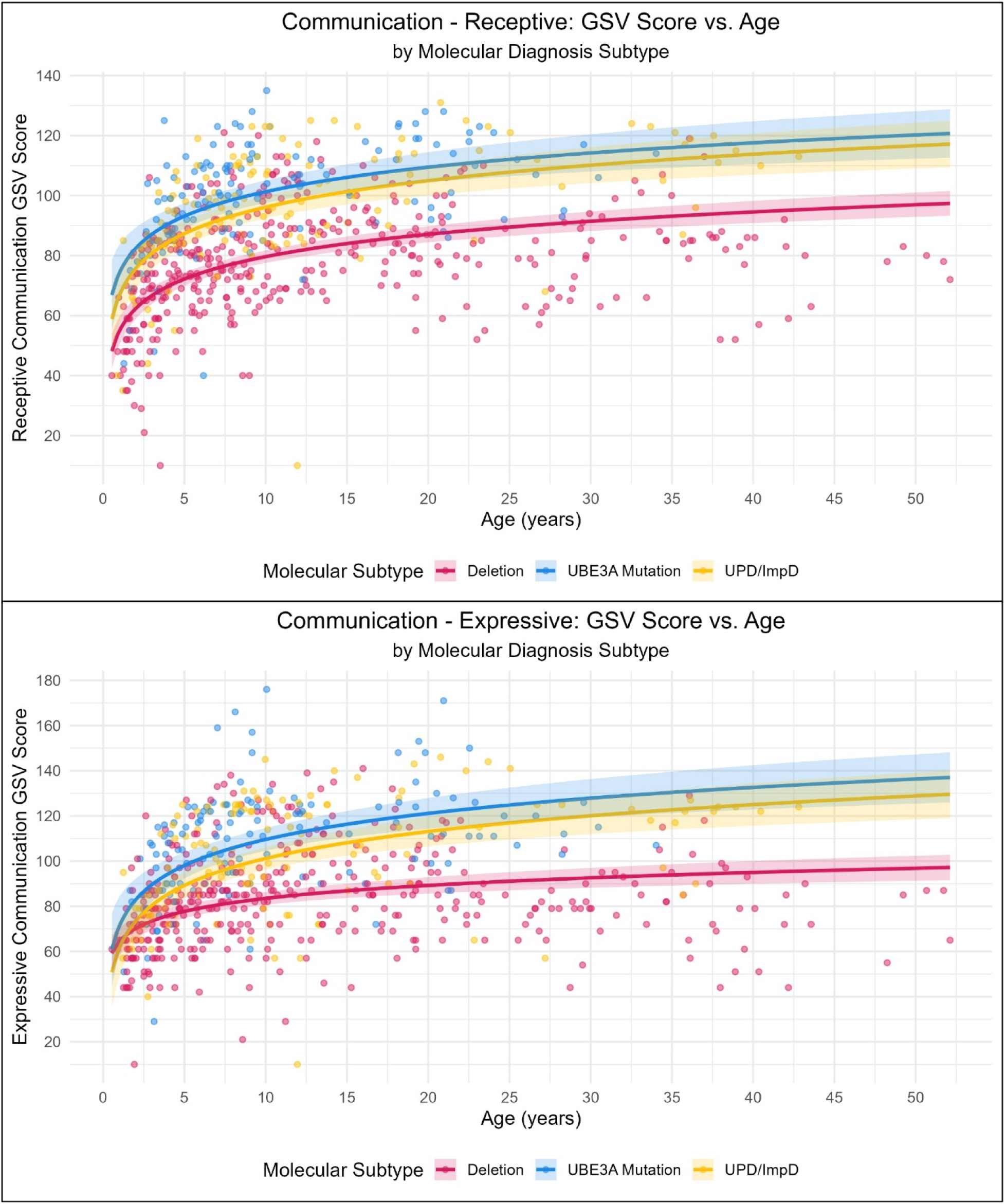
Effects plots of Vineland-3 growth scale values for the Communication domain by molecular subtype

**Table 3.** Model Estimates for Vineland-3 Communication Subdomain Growth Scale Values by Molecular Subtype.

|  | Communication |  |  |  |
| --- | --- | --- | --- | --- |
|  | Receptive |  | Expressive |  |
| Fixed Effects | $\beta$ (SE) | | $\beta$ (SE) | |
| Intercept | 80.66 (1.07) | *** | 84.27 (1.44) | *** |
| Age | 7.45 (0.69) | *** | 5.75 (0.94) | *** |
| Subtype |  |  |  |  |
| *UBE3A* mutation | 21.75 (2.23) | *** | 26.89 (2.99)<sup>^</sup> | *** |
| UPD/ImpD | 16.70 (2.21) | *** | 18.53 (2.97) | *** |
| Age × subtype |  |  |  |  |
| Age × *UBE3A* mutation | 0.70 (1.57) |  | 5.77 (2.16) | ** |
| Age × UPD/ImpD | 1.37 (1.51) |  | 6.17 (2.07) | ** |

*Note.* The deletion group served as the reference group. Age centered at 11 years of age and log<sub>10</sub>-transformed. UPD: uniparental disomy. ImpD: imprinting defect. <sup>^</sup>GSVs in the *UBE3A* mutation group were higher than those in the UPD/ImpD group ( $p < .05$ ).
\* $p < .05$ , \*\* $p < .01$ , \*\*\* $p < .001$

For Expressive Communication, a similar pattern was observed. Both the *UBE3A* mutation (β = 26.89, *SE* = 2.99, *p* < .001) and UPD/ImpD groups (β = 18.53, SE = 2.97, *p* < .001) had significantly higher GSVs at age 11 relative to the deletion group, and the UPD/ImpD group had significantly lower Expressive Communication GSVs at age 11 relative to the *UBE3A* mutation group (*p* = .023). Age was positively associated with Expressive Communication (β = 5.75, SE = 0.94, *p* < .001). However, unlike Receptive Communication, age-by-subtype interactions were significant for both the *UBE3A* mutation group (β = 5.77, SE = 2.16, *p* = .008) and the UPD/ImpD group (β = 6.17, SE = 2.07, *p* = .003), indicating slightly steeper age-related increases in Expressive Communication for these non-deletion subtypes relative to the deletion group. The trajectories of the non-deletion groups were not significantly different from each other (**Table 3**, **Figure 1**).

#### Daily Living Skills

For the Personal subdomain, both the *UBE3A* mutation (β = 18.78, *SE* = 1.93, *p* < .001) and UPD/ImpD groups (β = 13.04, *SE* = 1.92, *p* < .001) had significantly higher GSVs at age 11 relative to the deletion group. Additionally, the *UBE3A* mutation group had significantly higher GSVs at age 11 relative to the UPD/ImpD group (*p* = .016). Age was positively associated with Personal skills (β = 5.65, *SE* = 0.61, *p* < .001). Additionally, the age-by-subtype interaction was significant for the *UBE3A* mutation group (β = 4.20, *SE* = 1.39, *p* = .003), but not the UPD/ImpD group (β = 2.60, *SE* = 1.33, *p* = .052), signifying steeper age-related increases in Personal skills for the *UBE3A* mutation group relative to the deletion group. Despite this difference, the trajectories of the non-deletion groups were not significantly different from each other (**Table 4**, **Figure 2**).

**Figure 2.**
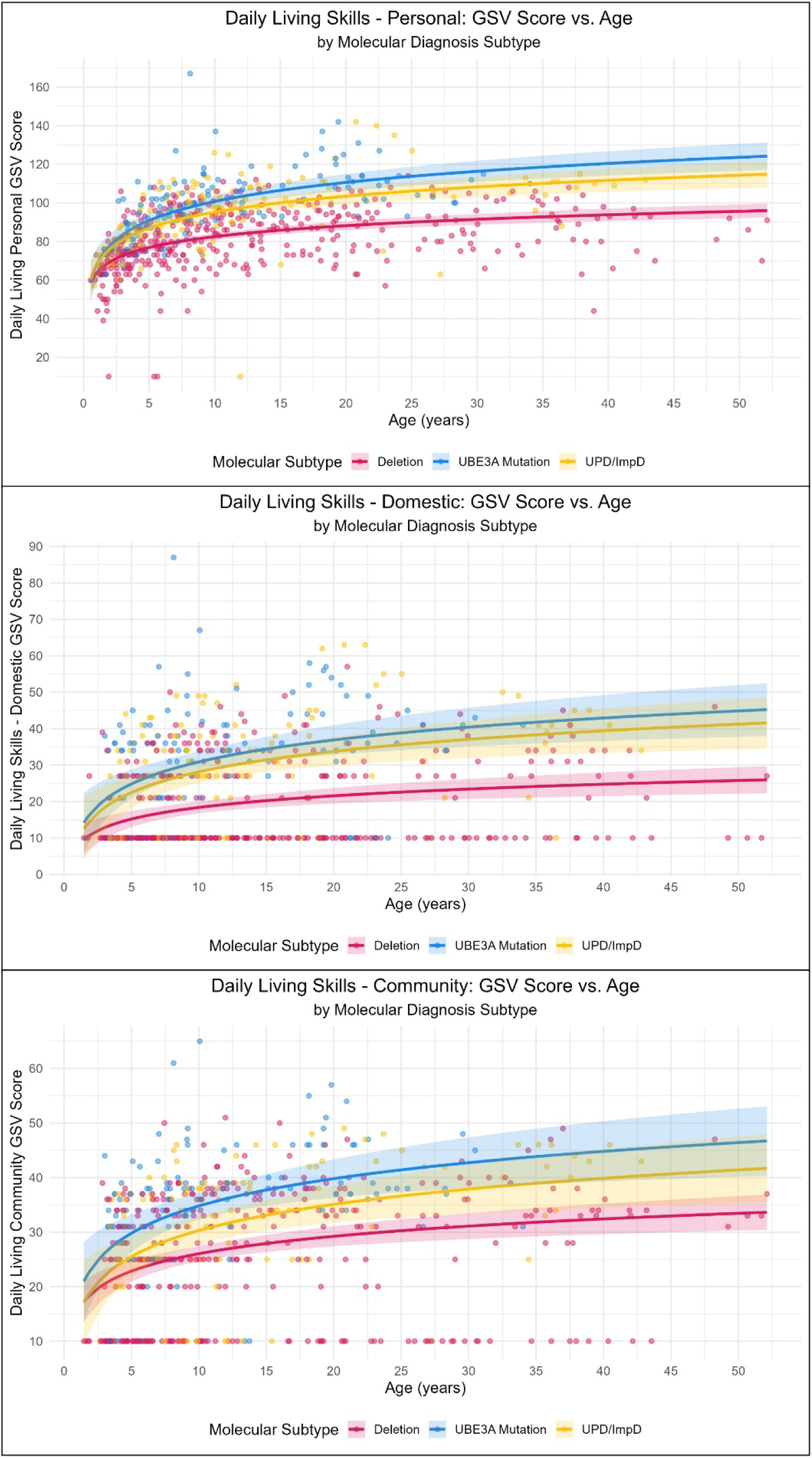
Effects plots of Vineland-3 growth scale values for the Daily Living Skills domain by molecular subtype

**Table 4.** Model Estimates for Vineland-3 Daily Living Skills Subdomain Growth Scale Values by Molecular Subtype.

| **Fixed Effects** | **Daily Living Skills** |  |  |  |  |  |
| --- | --- | --- | --- | --- | --- | --- |
|  | **Personal** |  | **Domestic** |  | **Community** |  |
|  | **$\beta$ (SE)** |  | **$\beta$ (SE)** |  | **$\beta$ (SE)** |  |
| Intercept | 83.30 (0.93) | *** | 18.81 (0.84) | *** | 26.48 (0.72) | *** |
| Age | 5.65 (0.61) | *** | 3.19 (0.73) | *** | 3.19 (0.64) | *** |
| Subtype |  |  |  |  |  |  |
| *UBE3A* mutation | 18.78 (1.93)<sup>^</sup> | *** | 12.88 (1.72) | *** | 9.01 (1.48)<sup>^</sup> | *** |
| UPD/ImpD | 13.04 (1.92) | *** | 10.09 (1.73) | *** | 4.48 (1.49) | ** |
| Age × subtype |  |  |  |  |  |  |
| Age × *UBE3A* mutation | 4.20 (1.39) | ** | 2.85 (1.59) |  | 1.81 (1.39) |  |
| Age × UPD/ImpD | 2.60 (1.33) |  | 2.46 (1.62) |  | 1.60 (1.42) |  |

*Note.* The deletion group served as the reference group. Age centered at 11 years of age and log<sub>10</sub>-transformed. UPD: uniparental disomy. ImpD: imprinting defect. <sup>^</sup>GSVs in *UBE3A* mutation group higher than UPD/ImpD group ( $p < .05$ ).
\* $p < .05$ , \*\* $p < .01$ , \*\*\* $p < .001$

In the Domestic subdomain, the *UBE3A* mutation (β = 12.88, *SE* = 1.72, *p* < .001) and UPD/ImpD groups (β = 10.09, *SE* = 1.73, *p* < .001) exhibited significantly higher GSVs at age 11 relative to the deletion group. At age 11, the *UBE3A* mutation and UPD/ImpD groups did not differ from each other. Age was positively associated with Domestic GSVs (β = 3.19, *SE* = 0.73, *p* < .001); however, there were no significant differences in trajectories between any of the subtype groups (**Table 4**, **Figure 2**).

The Community subdomain largely mirrored the patterns observed in the Personal subdomain, with both the *UBE3A* mutation (β = 9.01, *SE* = 1.48, *p* < .001) and UPD/ImpD groups (β = 4.48, *SE* = 1.49, *p* = .003) exhibiting higher GSVs at age 11 compared to the deletion group, and the *UBE3A* mutation group having higher GSVs relative to the UPD/ImpD group (*p* = .014). Age was positively associated with Community GSVs (β = 3.19, *SE* = 0.64, *p* < .001), but age-by-subtype interactions were not significant, indicating similar age-related trajectories across subtypes (**Table 4**, **Figure 2**).

#### Socialization

For the Interpersonal Relationships subdomain, both non-deletion subtypes showed significantly higher GSVs than the deletion group at age 11: the *UBE3A* mutation group scored, on average, 13.14 points higher (*SE* = 1.67, *p* < .001), and the UPD/ImpD group scored 7.64 points higher (*SE* = 1.66, *p* < .001); the UPD/ImpD group had significantly lower GSVs relative to the *UBE3A* mutation group (*p* = .008). Age was positively associated with Interpersonal Relationships GSVs (β = 3.92, *SE* = 0.53, *p* < .001), indicating that GSVs increased with age. However, the interaction effects of age-by-subtype were non-significant across all subtype group comparisons, indicating similar age-related trajectories across subtypes (**Table 5**, **Figure 3**).

**Figure 3.**
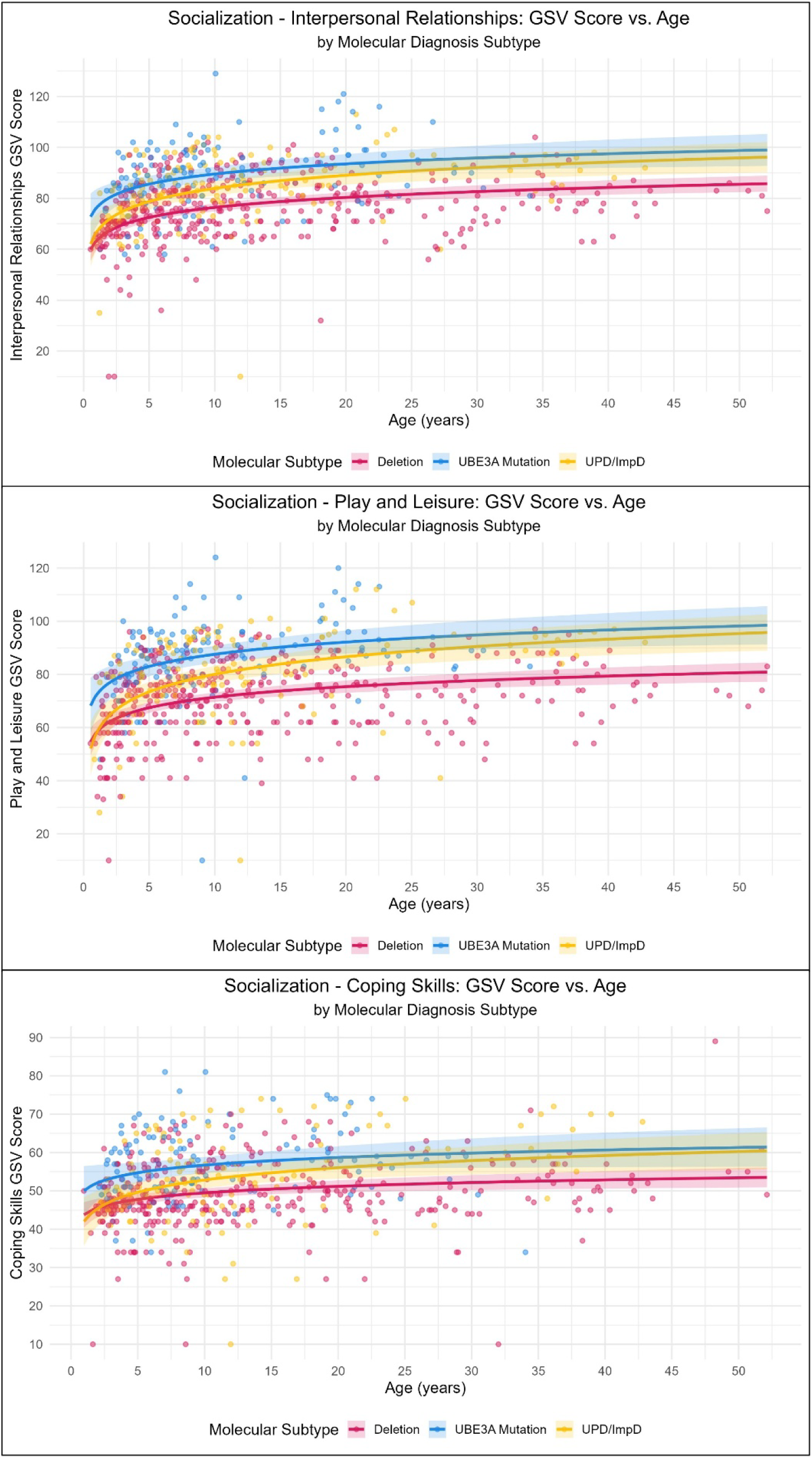
Effects plots of Vineland-3 growth scale values for the Socialization domain by molecular subtype

**Table 5.**
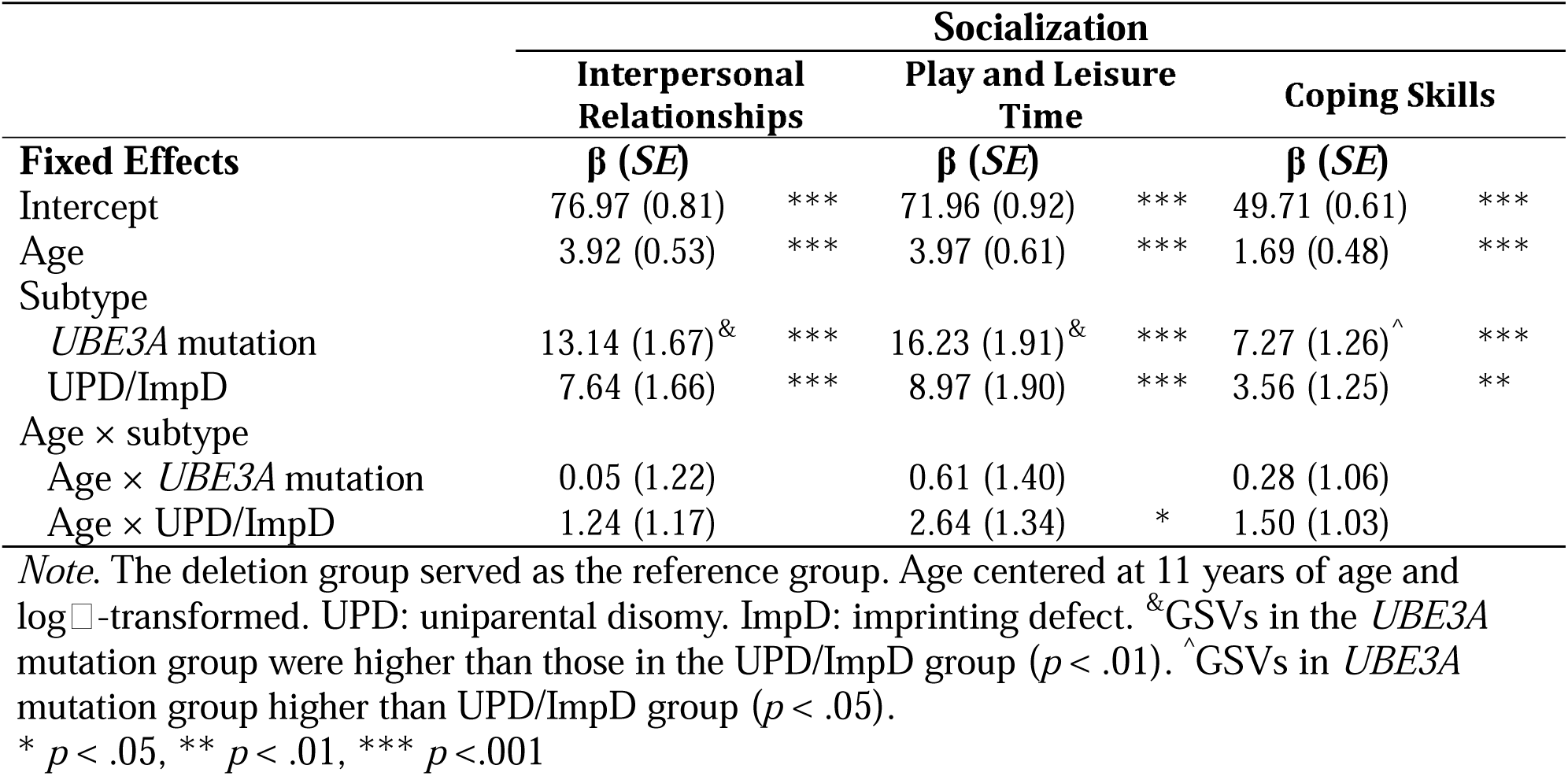
Model Estimates for Vineland-3 Socialization Subdomain Growth Scale Values by Molecular Subtype.

|  | Socialization |  |  |  |  |  |
| --- | --- | --- | --- | --- | --- | --- |
|  | Interpersonal Relationships |  | Play and Leisure Time |  | Coping Skills |  |
| Fixed Effects | $\beta$ (SE) | | $\beta$ (SE) | | $\beta$ (SE) | |
| Intercept | 76.97 (0.81) | *** | 71.96 (0.92) | *** | 49.71 (0.61) | *** |
| Age | 3.92 (0.53) | *** | 3.97 (0.61) | *** | 1.69 (0.48) | *** |
| Subtype |  |  |  |  |  |  |
| *UBE3A* mutation | 13.14 (1.67)<sup>&</sup> | *** | 16.23 (1.91)<sup>&</sup> | *** | 7.27 (1.26)<sup>^</sup> | *** |
| UPD/ImpD | 7.64 (1.66) | *** | 8.97 (1.90) | *** | 3.56 (1.25) | ** |
| Age × subtype |  |  |  |  |  |  |
| Age × *UBE3A* mutation | 0.05 (1.22) |  | 0.61 (1.40) |  | 0.28 (1.06) |  |
| Age × UPD/ImpD | 1.24 (1.17) |  | 2.64 (1.34) | * | 1.50 (1.03) |  |

*Note.* The deletion group served as the reference group. Age centered at 11 years of age and log<sub>10</sub>-transformed. UPD: uniparental disomy. ImpD: imprinting defect. <sup>&</sup>GSVs in the *UBE3A* mutation group were higher than those in the UPD/ImpD group ( $p < .01$ ). <sup>^</sup>GSVs in *UBE3A* mutation group higher than UPD/ImpD group ( $p < .05$ ).
\* $p < .05$ , \*\* $p < .01$ , \*\*\* $p < .001$

For the Play and Leisure Time subdomain, the *UBE3A* mutation (β = 16.23, *SE* = 1.91, *p* < .001) and UPD/ImpD groups (β = 8.97, *SE* = 1.90, *p* < .001) exhibited significantly higher GSVs at age 11 relative to the deletion group; again, the UPD/ImpD group had significantly lower GSVs relative to the *UBE3A* mutation group (*p* = .002). Age was positively associated with Play and Leisure Time GSVs (β = 3.97, *SE* = 0.61, *p* < .001), and the age-by-subtype interaction was significant for the UPD/ImpD group (β = 2.64, *SE* = 1.34, *p* = .049), but not the *UBE3A* mutation group (β = 0.61, *SE* = 1.40, *p* = .664). This finding indicates steeper age-related increases in Play and Leisure Time skills for the UPD/ImpD group relative to the deletion group. Trajectories between the UPD/ImpD and *UBE3A* mutation groups, however, were not significantly different from each other (**Table 5**, **Figure 3**).

Results for the Coping Skills subdomain were very similar to the patterns observed in the Interpersonal Relationships subdomain, with both the *UBE3A* mutation (β = 7.27, *SE* = 1.26, *p* < .001) and UPD/ImpD groups (β = 3.56, *SE* = 1.25, *p* = .005) exhibiting higher GSVs at age 11 compared to the deletion group. GSVs in the *UBE3A* mutation group were also significantly higher at age 11 than those in the UPD/ImpD group (*p* = .018). Age was positively associated with Coping Skills GSVs (β = 1.69, *SE* = 0.48, *p* < .001); however, age-by-subtype interactions were not significant across all subtype comparisons, indicating similar age-related trajectories across subtypes (**Table 5**, **Figure 3**).

#### Motor Skills

For the Gross Motor subdomain, both non-deletion subtypes showed significantly higher GSVs than the deletion group at age 11: the *UBE3A* mutation group scored, on average, 22.04 points higher (*SE* = 3.44, *p* < .001), and the UPD/ImpD group scored 15.18 points higher (*SE* = 3.45, *p* < .001). GSVs between the non-deletion groups at age 11 were not significantly different from each other. Age was positively associated with Gross Motor GSVs (β = 13.71, *SE* = 1.02, *p* < .001). However, the interaction effects of age-by-subtype were non-significant across all group comparisons, indicating similar age-related trajectories across subtypes (**Table 6**, **Figure 4**).

**Figure 4.**
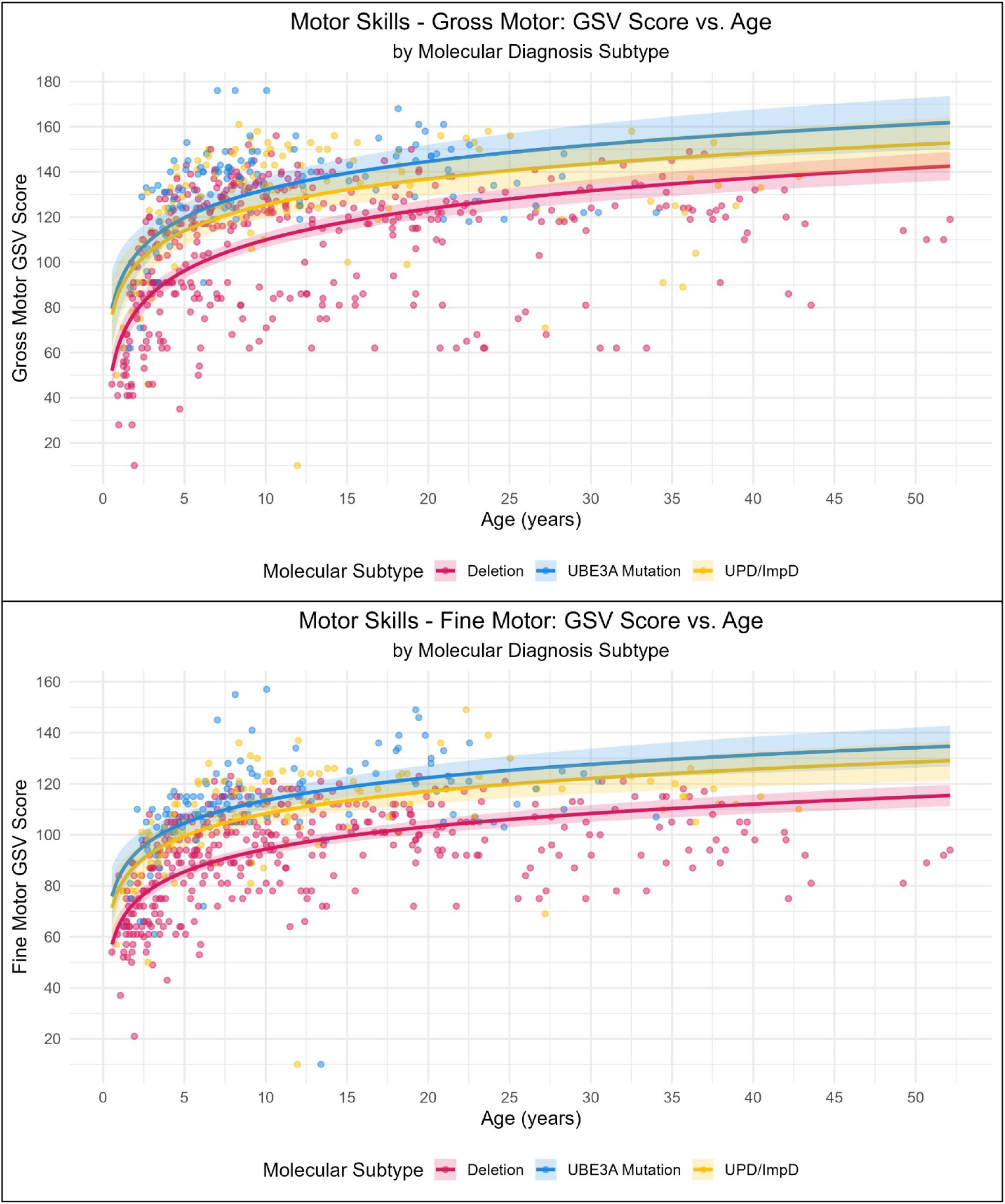
Effects plots of Vineland-3 growth scale values for the Motor Skills domain by molecular subtype

**Table 6.** Model Estimates for Vineland-3 Motor Skills Subdomain Growth Scale Values by Molecular Subtype.

| Fixed Effects | Motor Skills |  |  |  |
| --- | --- | --- | --- | --- |
| | Gross Motor<br>$\beta$ (SE) | | Fine Motor<br>$\beta$ (SE) | |
| Intercept | 111.78 (1.68) | *** | 95.55 (1.07) | *** |
| Age | 13.71 (1.02) | *** | 8.84 (0.69) | *** |
| Subtype |  |  |  |  |
| *UBE3A* mutation | 22.04 (3.44) | *** | 19.14 (2.19) | *** |
| UPD/ImpD | 15.18 (3.45) | *** | 13.92 (2.20) | *** |
| Age × subtype | | | | |
| Age × *UBE3A* mutation | -1.25 (2.26) | | 0.08 (1.57) | |
| Age × UPD/ImpD | -2.22 (2.20) | | -0.11 (1.51) | |

*Note.* The deletion group served as the reference group. Age centered at 11 years of age and log<sub>10</sub>-transformed. UPD: uniparental disomy. ImpD: imprinting defect.
\* $p < .05$ , \*\* $p < .01$ , \*\*\* $p < .001$

The Fine Motor subdomain mirrored the patterns observed in the Gross Motor subdomain, with both the *UBE3A* mutation (β = 19.14, *SE* = 2.19, *p* < .001) and UPD/ImpD groups (β = 13.92, *SE* = 2.20, *p* < .001) exhibiting higher GSVs at age 11 compared to the deletion group. These non-deletion groups, however, did not differ significantly from each other. Age was positively associated with Fine Motor GSVs (β = 8.84, *SE*= 0.69, *p* < .001), but age-by-subtype interactions were not significant, indicating similar age-related trajectories across all subtypes (**Table 6**, **Figure 4**).

## Discussion

In this large longitudinal cohort of individuals with AS, we characterized adaptive functioning trajectories using the Vineland-3, examining differences across molecular subtypes. Consistent with prior studies using the Vineland-II (e.g., Gentile et al., 2010; Gwaltney et al., 2024; Keute et al., 2021), individuals with deletion subtypes consistently demonstrated significantly lower adaptive functioning in all domains on the Vineland-3 compared to those with non-deletion subtypes. Across the Vineland-3, a clear genotype–phenotype gradient was evident: individuals with deletion subtype showed the greatest impairment in adaptive functioning, whereas individuals with non-deletion subtypes showed higher adaptive functioning overall; within the non-deletion group, individuals with UPD/ImpD tended to show more impairment (i.e., lower Vineland scores) than those with *UBE3A* point mutations, consistent with our baseline AE comparisons (particularly within Socialization and select Daily Living Skills and Communication outcomes). This pattern is biologically plausible given the relative extent of genetic disruption across subtypes: deletions remove the maternal *UBE3A* allele and additionally affect multiple genes within 15q11–q13, contributing to broader neurodevelopmental impact, whereas *UBE3A* point mutations primarily disrupt *UBE3A* itself. UPD/ImpD represents a distinct mechanism in which the individual effectively has two paternal copies (and no functional maternal contribution) for the imprinted 15q11–q13 region; thus, maternally expressed genes such as neuronal *UBE3A* remain silenced, and paternally expressed genes are present in a “double paternal” epigenetic context.

We also found that adaptive skills increased with age in all domains, though trajectories were nonlinear and varied modestly by subtype in some subdomains. The association between age and GSVs was nonlinear, best modeled using a log_2_ transformation of age, indicating that skill acquisition occurs more rapidly earlier in life and slows over time, consistent with clinical observations and past research in AS (Keute et al., 2021). The continued growth observed across domains suggests that adaptive skills remain malleable beyond early childhood. However, the slowing rate of growth may have implications for trial design, including expectations regarding the magnitude and timing of measurable change.

A major motivation for the present study was to evaluate adaptive functioning using the Vineland-3, which includes more items assessing early emerging skills on some subdomains and incorporates AAC modalities in the Communication domain. In a prior study using the Vineland-II, a substantial proportion of participants demonstrated floor effects in certain subdomains, limiting interpretability of AE scores (Gwaltney et al., 2024). The Vineland-3 appears to offer improved sensitivity through additional items at the lower end of the ability range for some subdomains. Moreover, the Vineland-3 offers improved assessment of communication abilities by capturing a broader range of modalities, including those beyond gestures, sounds, and spoken language by accounting for AAC use based on updated guidance from the publisher. This expanded scope allows for a more precise evaluation of functional communication skills.

Notably, the current study revealed differences in the slope by molecular subtype within the Expressive domain, whereas such differences were not observed in a previous study using the Vineland-II (Gwaltney et al., 2024). These findings highlight the advantage of Vineland-3 relative to the Vineland-II in detecting differences in expressive communication across subtypes. Nevertheless, floor-related challenges have not been entirely eliminated. Certain subdomains still reflect very limited skill acquisition in some individuals, particularly those with deletion subtypes. Gwaltney and colleagues previously reported that approximately 37% of observations were at the floor on the Domestic subdomain of the Vineland-II (Gwaltney et al., 2024). In the current study, the proportion at floor increased to nearly 46% (265 observations). We hypothesize that this increase reflects differences in item content—specifically, the inclusion of higher-level skills in the first few items on this subdomain in the Vineland-3 compared to the Vineland-II—as well as changes in scoring guidance that may have led to lower scores on the Vineland-3. In contrast, floor effects on the Community subdomain were comparable across studies (21% in the current sample vs. approximately 24% previously) (Gwaltney et al., 2024).

Importantly, floor-related interpretability challenges are further compounded by the structure of AE scores. As with the Vineland-II, many subdomains on the Vineland-3 have a range of raw scores at the low end of the scale that are associated with the same AE score (e.g., raw scores between 0 and 4 are associated with an AE score of 0:0 on the Expressive subdomain; raw scores between 0 and 23 are associated with an AE score of <2:0 on the Coping Skills subdomain). In our sample, where a substantial proportion of observations clustered at or near the lowest attainable raw scores in several subdomains, this many-to-one mapping can obscure meaningful within-person change and reduce sensitivity to detect growth, because increases in raw scores may not translate into changes in AE values. Raw scores avoid this specific compression, but they remain difficult to interpret developmentally across a wide age range and are not designed to function as an equal-interval metric. By contrast, GSVs are intended to provide a more continuous, approximately equal-interval scale that can support longitudinal modeling of growth, including in the presence of floor effects. Therefore, consistent with recent work in autistic children (Kwok et al., 2022) and children with rare pediatric neurodegenerative diseases (Eisengart et al., 2022), we used GSVs as our primary outcome for longitudinal analyses in AS. Their scaling properties can facilitate estimation of within-person change over time and model-based comparisons of trajectories within subdomains (Farmer et al., 2025; Farmer et al., 2023; Psimas & Williams, 2024).

Relative to previous work using the Vineland-II, the present findings reinforce core genotype–phenotype differences while extending prior natural history estimates with subdomain-specific, model-based trajectories on the Vineland-3 across a broader age range. The overall pattern—lower adaptive functioning in those with deletion subtypes compared to those with non-deletion subtypes, and relative weaknesses in expressive communication—remains consistent. In addition, the Vineland-3 enabled detection of modest subtype-related differences in age-associated change for Expressive Communication that were not observed in prior Vineland-II analyses (Gwaltney et al., 2024). The expanded item pool and alternative scoring framework in the Vineland-3 appear to improve detection of incremental growth, particularly at older ages and among individuals with lower levels of functioning. Importantly, the ability to model nonlinear trajectories across a wide age range, including adulthood, represents an advancement over prior analyses restricted to childhood and early adolescence. These data provide updated natural history benchmarks using an instrument that has been, and continues to be, increasingly deployed in clinical trials in AS. Notably, comparable GSV trajectories were recently reported in a natural history comparison sample of children aged 1–16 years that included participants with both deletion and *UBE3A* mutation subtypes and was used in the Phase 1 clinical trial of the *UBE3A*-ATS antisense oligonucleotide rugonersen (Hipp et al., 2025).

Although participants ranged in age up to 52 years, adult participants were underrepresented in the sample compared to children. As disease-modifying therapies continue to be developed, understanding adaptive trajectories across the full lifespan becomes increasingly important. More longitudinal data in adults will help determine whether adaptive growth plateaus, continues at a slow rate, or changes direction over time. Such data will also inform realistic outcome expectations for trials enrolling adolescents and adults.

### Limitations and Future Directions

Several limitations warrant consideration. First, although longitudinal, the number of study visits per participant was modest (mean of two visits), which limits precision in estimating within-person change. Second, while GSVs appear to provide advantages for modeling growth relative to score types that are vulnerable to floor effects (i.e., age-normed scores), they are not comparable across subdomains, restricting cross-domain interpretation of magnitude. Third, GSVs are derived from item response theory calibrations based on the Vineland-3 normative sample; although they are intended to function as approximately equal-interval measures, it is not guaranteed that the same item parameters and scaling properties generalize fully to AS. For example, the relative ordering of item difficulty and item discrimination could differ in AS (or show differential item functioning), which could alter the measurement properties of the GSV transformation in this population. Future work should evaluate these assumptions directly (e.g., by testing differential item functioning and/or estimating AS-specific calibrations where sample sizes permit). Fourth, as with all caregiver-report measures, responses on the Vineland-3 may be influenced by reporting and recall bias (Godoy et al., 2019; Khare & Vedel, 2019; Zheng et al., 2024).

Additionally, the modeling approach using linear mixed effects models and log_2_(age) captures the data better than linear age, and visual inspections suggest an overall good fit (see figures). Future work with more data may test refined non-linear relationships with age and consider modeling approaches that do not assume homoscedasticity and normally distributed errors.

Future research would benefit from incorporating additional longitudinal assessments, especially across adolescence and adulthood. Sustained follow-up into adulthood will be critical for understanding developmental trajectories across domains of adaptive functioning. Increasing diversity in terms of race and ethnicity would also benefit future studies. Additionally, evaluating the correspondence between Vineland-3 scores and real-world functioning captured via video-based assessment is an important next step, particularly as low-burden, remote assessment tools are increasingly incorporated into clinical research in AS (Leffler et al., 2026). As the Vineland-3 is increasingly used as a clinical trial endpoint in AS, robust natural history data such as these are essential for contextualizing treatment-related change and for guiding power calculations, endpoint selection, and interpretation of clinical meaningfulness.

## Conclusions

Adaptive functioning in individuals with AS improves across childhood and into adulthood, though levels vary by molecular subtype, with individuals with deletion subtypes showing consistently lower skills relative to those with non-deletion subtypes. Expressive communication remains a relative area of weakness across groups, even after inclusion of AAC guidance in the Vineland-3. Given known floor effects and AE score compression in AS, GSVs provide a practical, growth-oriented metric for modeling longitudinal change on the Vineland-3, and in the present analyses they supported estimation of nonlinear trajectories across subdomains despite substantial floor effects in select areas. As the Vineland-3 is increasingly used as a clinical trial endpoint in AS, careful selection of scoring metrics (including consideration of GSVs alongside raw score analyses) will be important for maximizing interpretability and sensitivity to change. These findings provide updated natural history benchmarks to guide intervention planning, trial design, and realistic expectations for individuals with AS.

## Data Availability

The data sets used and/or analyzed during this study are available from SNP on reasonable request.

