## Supplementary material for "Assessment of adaptive functioning in Angelman syndrome using the Vineland Adaptive Behavior Scales, Third Edition"

**Supplemental Table 1**

*Comparison of Model Metrics for Receptive Communication GSV Scores*

| **Model** | **AIC** | **BIC** | **Marginal R^2^** |
| --- | --- | --- | --- |
| Spline | 5293.4 | 5324.8 | 0.235 |
| Linear | 5453.9 | 5471.9 | 0.119 |
| Quadratic | 5384.8 | 5407.2 | 0.219 |
| Log(age) | 5347.6 | 5365.5 | 0.247 |
| Log_2_(age) | 5348.3 | 5366.3 | 0.247 |
| Log_2_(age) + Log(GSV score) | N/A | N/A | 0.225 |

*Note*. The AIC and BIC values for the model with Log_2_(age) + Log(score) is not comparable to the AIC and BIC values for the other models because *y* is logged.

**Supplemental Table 2**

*Model Estimates for Vineland-3 Communication Subdomain Raw Scores by Molecular Subtype*

|  | **Communication** | | | |
| --- | --- | --- | --- | --- |
|  | **Receptive** | | **Expressive** | |
| **Fixed Effects** | **β (*SE*)** |  | **β (*SE*)** |  |
| Intercept | 27.55 (0.87) | *** | 16.30 (0.81) | *** |
| Age | 5.32 (0.55) | *** | 2.34 (0.52) | *** |
| Subtype |  |  |  |  |
| *UBE3A* mutation | 19.27 (1.80) | *** | 15.04 (1.69) | *** |
| UPD/ImpD | 15.09 (1.78) | *** | 9.95 (1.68) | *** |
| Age × subtype |  |  |  |  |
| Age × *UBE3A* mutation | 2.08 (1.26) |  | 4.55 (1.19) | *** |
| Age × UPD/ImpD | 2.15 (1.21) |  | 4.70 (1.14) | *** |

*Note*. The deletion group served as the reference group. Age centered at 11 years of age and log₂-transformed. UPD: uniparental disomy. ImpD: imprinting defect.

*** *p* <.001

**Supplemental Table 3**

*Model Estimates for Vineland-3 Daily Living Skills Subdomain Raw Scores by Molecular Subtype*

|  | **Daily Living Skills** | | | | | |
| --- | --- | --- | --- | --- | --- | --- |
|  | **Personal** | | **Domestic** | | **Community** | |
| **Fixed Effects** | **β (*SE*)** |  | **β (*SE*)** |  | **β (*SE*)** |  |
| Intercept | 21.87 (0.85) | *** | 1.68 (0.34) | *** | 4.32 (0.34) | *** |
| Age | 3.97 (0.53) | *** | 0.80 (0.29) | ** | 1.26 (0.29) | *** |
| Subtype |  |  |  |  |  |  |
| *UBE3A* mutation | 18.73 (1.76) | *** | 4.67 (0.70) | *** | 4.69 (0.69) | *** |
| UPD/ImpD | 13.05 (1.92) | *** | 3.41 (0.70) | *** | 2.05 (0.69) | ** |
| Age × subtype |  |  |  |  |  |  |
| Age × *UBE3A* mutation | 6.43 (1.19) | *** | 1.29 (0.04) | * | 1.51 (0.64) | * |
| Age × UPD/ImpD | 4.56 (1.15) | *** | 0.96 (0.14) |  | 1.48 (0.65) | * |

*Note*. The deletion group served as the reference group. Age centered at 11 years of age and log₂-transformed. UPD: uniparental disomy. ImpD: imprinting defect.

* *p* < .05, ** *p* < .01, *** *p* <.001

**Supplemental Table 4**

*Model Estimates for Vineland-3 Socialization Subdomain Raw Scores by Molecular Subtype*

|  | **Socialization** | | | | | |
| --- | --- | --- | --- | --- | --- | --- |
|  | **Interpersonal Relationships** | | **Play and Leisure Time** | | **Coping Skills** | |
| **Fixed Effects** | **β (*SE*)** |  | **β (*SE*)** |  | **β (*SE*)** |  |
| Intercept | 29.09 (0.65) | *** | 14.60 (0.60) | *** | 15.68 (0.60) | *** |
| Age | 2.71 (0.43) | *** | 2.04 (0.39) | *** | 1.64 (0.46) | *** |
| Subtype |  |  |  |  |  |  |
| *UBE3A* mutation | 11.89 (1.35) | *** | 13.06 (1.25) | *** | 7.80 (1.24) | *** |
| UPD/ImpD | 6.63 (1.34) | *** | 7.47 (1.24) | *** | 4.43 (1.23) | *** |
| Age × subtype |  |  |  |  |  |  |
| Age × *UBE3A* mutation | 1.32 (0.99) |  | 1.80 (0.89) | * | 0.64 (1.04) |  |
| Age × UPD/ImpD | 1.60 (0.95) |  | 3.07 (0.85) | *** | 2.35 (1.00) | * |

*Note*. The deletion group served as the reference group. Age centered at 11 years of age and log₂-transformed. UPD: uniparental disomy. ImpD: imprinting defect.

* *p* < .05, *** *p* <.001

**Supplemental Table 5**

*Model Estimates for Vineland-3 Motor Skills Subdomain Raw Scores by Molecular Subtype*

|  | **Motor Skills** | | | |
| --- | --- | --- | --- | --- |
|  | **Gross Motor** | | **Fine Motor** | |
| **Fixed Effects** | **β (*SE*)** |  | **β (*SE*)** |  |
| Intercept | 39.06 (1.18) | *** | 22.07 (0.51) | *** |
| Age | 7.59 (0.72) | *** | 3.59 (0.32) | *** |
| Subtype |  |  |  |  |
| *UBE3A* mutation | 18.21 (2.41) | *** | 10.90 (1.03) | *** |
| UPD/ImpD | 13.50 (2.42) | *** | 7.81 (1.04) | *** |
| Age × subtype |  |  |  |  |
| Age × *UBE3A* mutation | 2.50 (1.61) |  | 1.49 (0.72) | * |
| Age × UPD/ImpD | 1.35 (1.56) |  | 1.16 (0.69) |  |

*Note*. The deletion group served as the reference group. Age centered at 11 years of age and log₂-transformed. UPD: uniparental disomy. ImpD: imprinting defect.

* *p* < .05, *** *p* <.001
